# Comparison of a Novel Plethysmometric Method for Blood Pressure Estimation with Reference Sphygmomanometry

**DOI:** 10.1101/2025.11.15.25340287

**Authors:** Naveen Gangadharan, Bowya Baskaran, A Manavalan, M Praveen Kumar, M Anandhakumar, Benjamin Jebaraj, Suresh Devasahayam, Sathya Subramani

**Affiliations:** Christian Medical College, Thorapadi post, Vellore, 632002, Tamil Nadu, India, Affiliated to The TN MGR Medical University, Tamilnadu, India

**Keywords:** Plethysmometry, Sphygmomanometry, Hypertension, BP variability

## Abstract

**Background:** Plethysmometry is a cuff based method that records photoplethysmographic pulse amplitudes at multiple cuff pressure levels and estimates ranges of systolic and diastolic blood pressure together with mean arterial pressure.

**Objective:** To compare plethysmometric BP estimates with carefully obtained auscultatory sphygmomanometric measurements and to determine classification thresholds of plethysmometric pressures for hypertension diagnosis by comparison with sphygmomanometric reference.

**Methods:** Ninety six adults underwent four video and audio recorded mercury sphygmomanometric measurements (M0 to M4), two before and two after a single plethysmometric recording. Two observers independently interpreted the recordings, with adjudication when their readings differed by more than 2 mmHg. The mean of measurements M1 to M3 was used as the reference sphygmomanometric pressure.

Plethysmometry recorded cuff pressure and finger photoplethysmograms from the cuffed and uncuffed arms during 16 cuff pressure plateaus lasting for ten seconds each. From the relationship of pulse amplitude (in the cuffed arm) to cuff pressure during the plateaus, lower and upper limits of systolic and diastolic pressures were derived. Average systolic and diastolic pressures were calculated as midpoints of their ranges. Average pulse pressure was estimated. Mean arterial pressure (MAP) was estimated independently from cuff pressure oscillation amplitude; when the maximum cuff oscillation amplitude extended across adjacent pressure levels, lower and upper MAP estimates were reported and their midpoint used as the average MAP.

Agreement of plethysmometry and sphygmomanometry for average systolic and diastolic pressures was evaluated using mean differences, Bland Altman analysis, the numerical requirements of AAMI/ESH/ISO Criterion 1, and the historical BHS grading system. ROC analysis was used to explore plethysmometric thresholds for classifying hypertension, as compared to sphygmomanometric thresholds of 140 mmHg for systolic and 90 mmHg for diastolic pressures.

**Results:** The mean difference between plethysmometric and sphygmomanometric estimates was 0.97 [6.58] mmHg (mean[SD]) for systolic pressure and -3.02 [8.74] mmHg for diastolic pressure, meeting the numerical limits specified in AAMI/ESH/ISO Criterion 1 for systolic pressure, but not for diastolic pressure. Historical BHS grading was Grade A for systolic and Grade C for diastolic pressures.

Thirteen participants were hypertensive by the sphygmomanometric method, using the conventional thresholds of 140 mmHg and 90 mmHg for systolic and diastolic respectively. In exploratory ROC analysis, a classification criterion of plethysmometric diastolic pressure ≥ 83 mmHg OR mean arterial pressure ≥ 98 mmHg provided the highest Youden index, with a sensitivity of 100% and specificity of 90.36% as compared to sphygmomanometric classification. However, in order not to miss isolated systolic hypertension, a criterion of (systolic ≥ 124 mmHg OR diastolic ≥ 83 mmHg) AND MAP ≥ 98 mmHg was employed in the study. Twelve of the thirteen hypertensives were identified by this criterion, with one discordant negative. Five additional participants were classified as hypertensive by plethysmometry. The five discordant positives and one discordant negative by plethysmometry would require further investigations to confirm the classification.

**Conclusions:** A single plethysmometric recording showed small mean differences from the average of three reference sphygmomanometric measurements and promising discrimination of sphygmomanometrically defined hypertension. Because the complete AAMI/ESH/ISO validation protocol was not performed and the diagnostic threshold was derived and evaluated in the same small cohort, these findings require confirmation using repeated paired measurements and independent validation datasets.

## Introduction

Hypertension is the leading cause of premature death (1). A single clinic BP reading is insufficient to diagnose hypertension, owing to biological variability and the phenomena of white-coat hypertension (WCH) and masked hypertension (MH). Current guidelines recommend repeated measurements and emphasize out-of-office BP monitoring for identifying white-coat and masked hypertension (2). The out-of-office methods available are 24-hour ambulatory blood pressure monitoring (ABPM) and home-based BP measurement (HBPM), also referred to as self-measured BP. Although ABPM is clinically valuable, its use in routine Indian practice remains limited. Kaul *et al* report that most surveyed physicians used ABPM in fewer than 5% of patients and demonstrate substantial WCH and MH prevalence (3). The India Heart study found substantial disagreement between office and self-measured classifications (4).

We have developed a cuff-based method—**Plethysmometry**—which measures pulse-volume changes at 16 cuff-pressure levels over a 6-minute period. The relationship of pulse amplitude to cuff-pressure is sigmoid. Systolic and diastolic pressure ranges were derived algorithmically from the transition regions of the cuff-pressure–pulse-amplitude relationship, together with pulse presence or absence, absolute and reference-normalized pulse amplitudes, within-plateau variability of pulse amplitude and the independently estimated MAP. Our method reports the ranges of systolic and diastolic pressures over the 6-minute period. We hypothesised that the pressure estimates obtained during a single plethysmometric recording could provide BP classification comparable to that obtained from multiple conventional measurements.

In this study, 4 audio and video recorded sphygmomanometric measurements were performed with a minimum 5-minute interval between the measurements. Plethysmometric measurement was done between the second and the third sphygmomanometric measurement. Sphygmomanometric pressures were reported by 2 independent observers. When the two primary-observer estimates differed by 2 mmHg or less, their values were averaged. When they differed by more than 2 mmHg, a third observer adjudicated the measurement; the primary-observer value that agreed with the adjudicator was retained, and the final reading was calculated as the mean of the two concordant values.

Validation procedures for new BP devices are for devices reporting a single pressure value and there are no protocols for devices reporting a range. Our attempt in this exploratory study is to perform comparative analysis, taking cues from the recommendations of AAMI/ESH/ISO validation standard (5) and BHS protocol (6).

Existing device-validation protocols were developed primarily for devices reporting single systolic and diastolic values from repeated test measurements. Plethysmometry instead reports pressure ranges from a single recording intended to capture short-term variation. The present exploratory comparison therefore assessed both midpoint agreement between the plethysmometric and repeated sphygmomanometric pressure ranges, as well as diagnostic agreement for hypertension classification.

We compare the average of the systolic and diastolic ranges obtained by a single plethysmometric recording in 96 individuals with averages derived from the last three of the four video- and audio-documented sphygmomanometric measurements reported by 2 independent observers. The systolic mean difference and standard deviation met the numerical limits specified in AAMI/ESH/ISO Criterion 1, whereas the diastolic standard deviation did not. The distributions of absolute differences corresponded to historical BHS Grade A for systolic and Grade C for diastolic pressures.

In addition, Receiver-operating-characteristic analysis (ROC analysis) was performed for the plethysmometric values to obtain the best classification parameter that matches the diagnostic performance of plethysmometry as compared to reference sphygmomanometry.

Even when a device meets the AAMI and BHS validation criteria, its diagnostic accuracy at the individual level cannot be assumed. The AAMI criteria permit a standard deviation of up to 8 mmHg, while the BHS criteria allow a proportion of readings to differ by more than 15 mmHg. Thus, although overall device performance may be acceptable at the population level, individual readings may still falsely identify or fail to detect hypertension. As with any diagnostic method, BP devices should therefore also be evaluated specifically for diagnostic sensitivity and specificity.

The physiological basis, instrumentation and complete analytical approach are described in an accompanying paper. The present study evaluates numerical agreement with reference sphygmomanometry and explores the diagnostic discrimination of plethysmometric parameters.

## Methods

This study was approved by the Institutional Review Board of Christian Medical College, Vellore, India (IRB Minute No. 14388, dated 22 December 2021). Adults aged ≥ 18 years were recruited from the local community between 1 March and 24 June 2022 after providing written informed consent. The prespecified exclusion criterion was the presence of an arrhythmia; however, no enrolled participant met this criterion. Of 100 participants enrolled, 96 had complete paired plethysmometric and sphygmomanometric data and were included in the final analysis. No participant was excluded after examination of the reference BP values.

### Reference Sphygmomanometry

Each participant underwent four mercury sphygmomanometric measurements, designated M0–M3, in a soundproof room. Two measurements were obtained before and two after plethysmometry, with at least five minutes between consecutive BP procedures; plethysmometry was therefore performed between M1 and M2. The mercury column was video-recorded while Korotkoff sounds were recorded simultaneously. Two observers independently interpreted each synchronized video–audio recording. When the two primary-observer estimates differed by 2 mmHg or less, their values were averaged. When they differed by more than 2 mmHg, a third observer adjudicated the measurement; the primary-observer value that agreed with the adjudicator was retained, and the final reading was calculated as the mean of those two concordant values.

### Plethysmometric measurement

Participants were seated in an armless chair with back support. Mid-arm circumference was measured, and an appropriately sized cuff was placed on one upper arm. Photoplethysmographic sensors were attached to corresponding fingers of the cuffed and contralateral arms, with the hands supported on the thighs and the palms facing upward.

The plethysmometric system simultaneously recorded cuff pressure and finger photoplethysmograms from the cuffed and uncuffed arms at a sampling frequency of 500 samples per second.

An initial cuff-pressure ramp was used to identify the approximate pressure at which the cuffed-arm pulse disappeared. Sixteen cuff-pressure levels, spanning 30 mmHg to approximately 30 mmHg above this preliminary systolic estimate, were then applied in descending order (The current protocol applies the same pressure levels in ascending order). Each pressure level was maintained for 10 seconds and was followed by complete cuff deflation for 10 seconds.

At each pressure plateau, cuffed-arm pulse amplitude was calculated from the pulse peak and preceding trough and was analysed both as an absolute value and after normalization to the simultaneously recorded contralateral-arm pulse amplitude. The relationship between pulse amplitude and cuff pressure was analysed algorithmically to derive lower and upper systolic and diastolic pressures. Mean arterial pressure was estimated independently from the cuff pressure at which oscillation amplitude was maximal. Full technical details of the device, recording protocol, and analytical algorithm are described in the accompanying paper (submitted to medRxiv).

The same prespecified, rule-based algorithm for analysis of the recorded signals was applied to all 96 participants. The analytical rules and algorithm parameters were finalized before the reference sphygmomanometric values from the present cohort were examined. The analytical algorithm was developed from the recurring cuff-pressure oscillation and PPG patterns observed in more than 1,000 recordings obtained in an ongoing study. Algorithm development was based on physiological signal features and did not use reference blood pressure values from those participants for calibration or optimization. The algorithm contained no machine-learning component.

Average plethysmometric systolic, diastolic and mean arterial pressures were calculated as the midpoints of the corresponding lower and upper estimates. Plethysmometric midpoint systolic and diastolic pressures were compared with the mean of sphygmomanometric measurements M1–M3. Bias and the standard deviation of the device–reference differences were calculated. The proportions of absolute differences within 5, 10 and 15 mmHg were also determined.

### Statistical analysis

Continuous variables were summarized using mean, standard deviation, median and interquartile range. Plethysmometric midpoint systolic and diastolic pressures were compared with the mean of reference measurements M1–M3. Differences were defined as plethysmometry minus reference sphygmomanometry. Agreement was assessed using mean difference, standard deviation of differences and Bland–Altman 95% limits of agreement (7). Proportional bias was assessed by linear regression of the device-reference difference against the mean of the two methods. The observed mean difference and standard deviation were compared with the numerical limits of AAMI/ESH/ISO Criterion 1 (5), and absolute differences were summarized using the historical BHS grading scheme (6).

Reference hypertension was defined as mean sphygmomanometric systolic pressure ≥140 mmHg or diastolic pressure ≥90 mmHg. ROC curves were generated separately for plethysmometric midpoint SBP, midpoint DBP and midpoint MAP. Candidate thresholds with both sensitivity and specificity exceeding 0.80 were examined. Candidate thresholds and combined classification criteria were derived and evaluated in the same cohort and therefore should be considered exploratory. Statistical analyses were performed using MATLAB R2018a.

## Results

The study subjects included 40 women and 56 men. Table 1 gives anthropometric data of the sample.

**Table 1.**
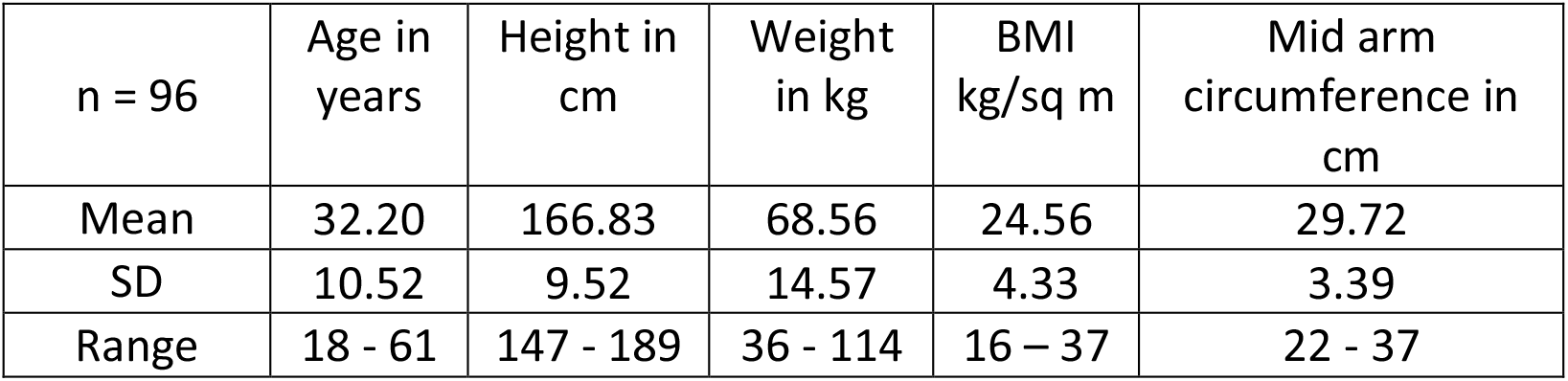
Anthropometric data of study subjects (n=96)

The range of plethysmometric pressures in each individual and the corresponding range of pressures from reference sphygmomanometric measurements M1 to M3 is shown in figure 1.

**Figure 1:**
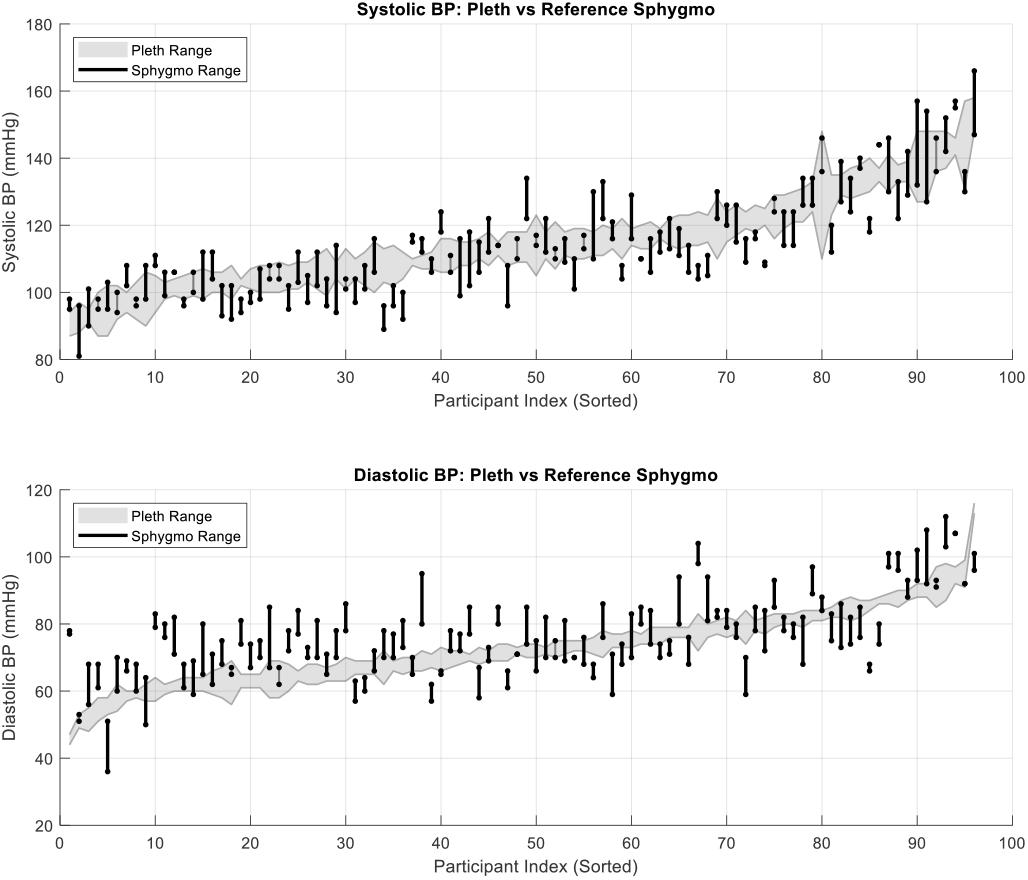
Plethysmometric and reference sphygmomanometric pressure ranges in individual participants – the grey band represents the plethysmometric range and the black vertical lines represent the range from three sphygmomanometric measurements. Participants were ordered by the average of plethysmometric systolic or diastolic ranges.

Variability of systolic and diastolic pressures as measured by Plethysmometry and 3 repeats of sphygmomanometry are given in figure 2:

**Figure 2:**
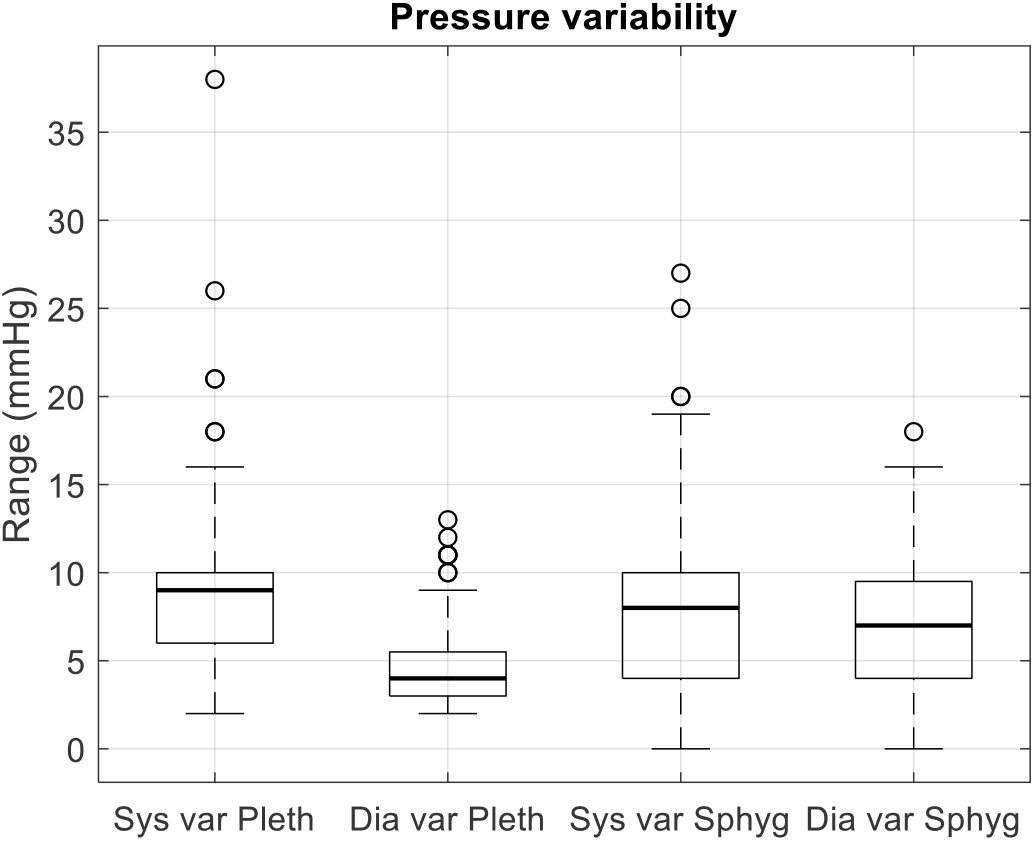
Interquartile ranges of systolic and diastolic variability as given by Plethysmometry and three repeats of sphygmomanometry (M1 to M3)

As there is no recommended format for comparison of pressure ranges, comparison of average pressures from both methods was performed.

### Average Pressures

Average systolic and diastolic pressures were computed as the within-subject mean of lower and upper limits for each individual, then aggregated across participants; the resulting group means for average systolic and diastolic pressures are shown as ‘Average systolic’ and ‘Average diastolic’ in the following tables.

### Agreement Between Methods

Differences were calculated as plethysmometry minus reference sphygmomanometry. Limits of agreement were calculated as the mean difference±1.96 standard deviations. Mean difference or Bias, SD, and limits of agreement for systolic and diastolic pressures are given in Table 2.

**Table 2:** Mean difference (Bias) and SD of differences (Plethysmometry – Reference sphygmomanometry) Bland–Altman analysis showed a mean plethysmometry–sphygmomanometry difference of ^0.97 ± 6.58^ mmHg for average systolic pressure and -3.02 ± 8.74 mmHg for diastolic pressure. 95% limits of agreement are shown in the table above.

| Pressure parameters in mmHg | Mean difference (Bias) | SD of differences | Lower limit of agreement | Upper limit of agreement |
| --- | --- | --- | --- | --- |
| Average systolic | 0.97 | 6.58 | -11.94 | 13.87 |
| Average diastolic | -3.02 | 8.74 | -20.15 | 14.11 |

### Comparison with the numerical limits of AAMI/ESH/ISO Criterion 1

According to the Association for the Advancement of Medical Instrumentation (AAMI) criteria for device validation, comparisons are to be made with the reference measurements done before and after each of the three test measurements. The absolute value of ‘bias’ (mean difference in the pressures computed for the whole sample) must be ≤ 5 mmHg and the SD of differences ≤ 8 mmHg. Since only one test measurement was performed in this case, the bias and SD are reported by comparing the midpoint of the systolic and diastolic range from plethysmometry with the average of the last three of the 4 reference sphygmomanometric measurements. This comparison with the numerical limits of Criterion 1 should not be interpreted as completion of the full AAMI/ESH/ISO validation protocol. With this limitation, it can be seen from Table 2 that the method will pass AAMI/ESH/ISO criterion 1 for systolic pressure, but not diastolic pressure.

Linear regression of the device–reference differences against the mean of the two methods demonstrated a small but significant proportional bias for systolic pressure (slope ™0.104, 95% CI ™0.201 to ™0.008; P = 0.035), with the difference between methods becoming progressively more negative as mean systolic pressure increased. There was no significant proportional bias for diastolic pressure (slope -0.036, P=0.672)

Bland-Altman plots with the mean of pressures from plethysmometry and sphygmomanometry on the X axis and the difference between the two methods on the Y axis, are given in Figure 3.

**Figure 3:**
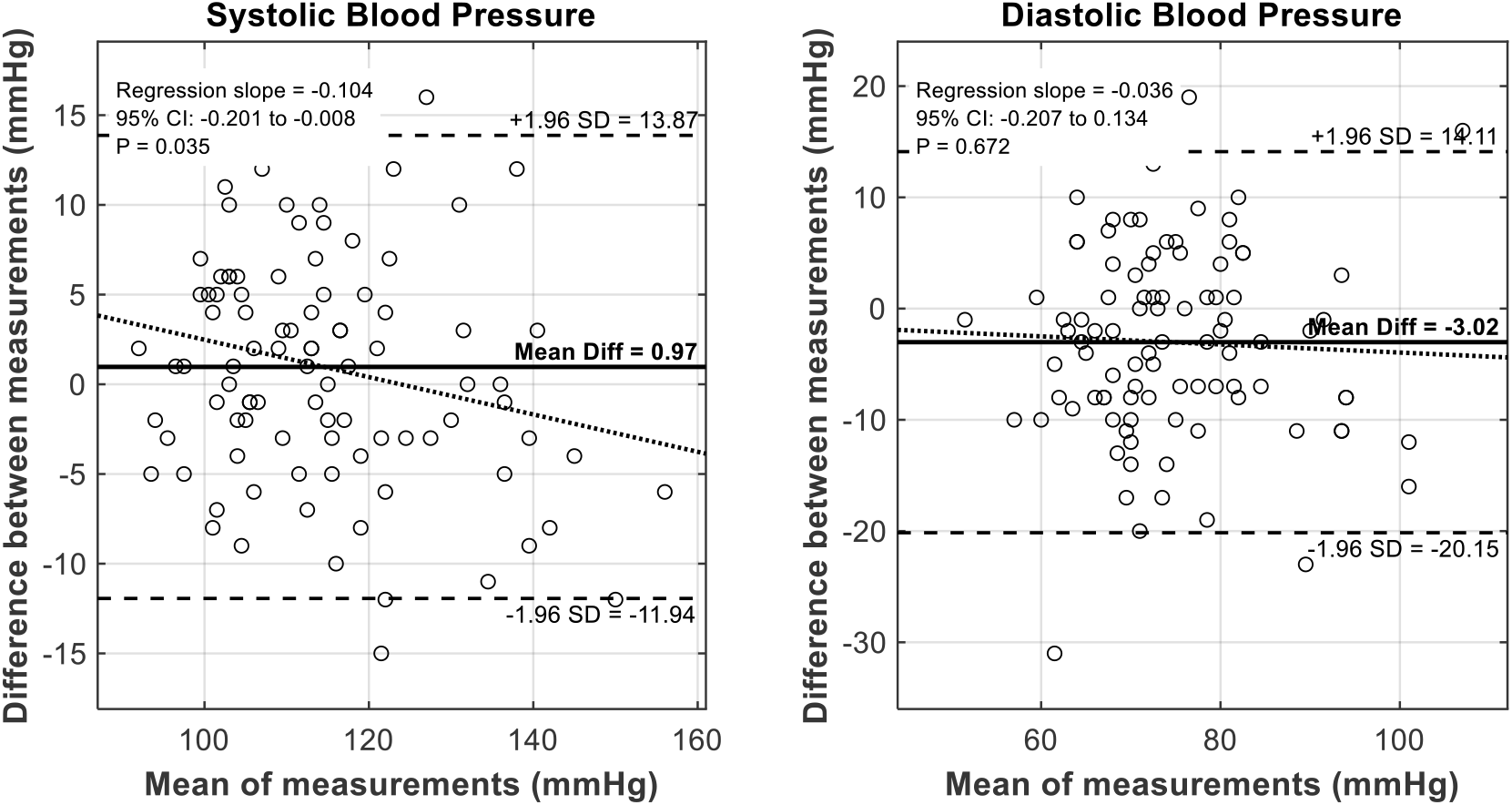
Bland-Altman plots for Plethysmometry versus Sphygmomanometry.

### BHS Grading

Under the historical BHS grading scheme, Grade A requires at least 60%, 85% and 95% of absolute device–reference differences to be within 5, 10 and 15 mmHg, respectively. Grade B requires at least 50%, 75% and 90%, and Grade C requires at least 40%, 65% and 85%; performance below Grade C is assigned Grade D. All three percentage thresholds must be met for assignment of a grade. The complete BHS protocol requires 85 participants and three device measurements per participant. In the present study, the percentage thresholds were applied descriptively to one plethysmometric midpoint– reference average difference per participant and should not be interpreted as completion of the formal BHS protocol.

When the observed absolute differences were assessed using the historical BHS percentage thresholds (Table 3), systolic pressure measurements attained Grade A, while diastolic pressure measurements were Grade C. Whether the same grades would be obtained under the complete BHS protocol, using three test-device measurements per participant, remains to be seen.

**Table 3:** BHS grading of agreement between plethysmometric and sphygmomanometric pressures.

| Difference between methods-> | ≤ 5 mmHg (%) | ≤ 10 mmHg (%) | ≤ 15 mmHg (%) | Grade |
| --- | --- | --- | --- | --- |
| Average systolic | 60.42 | 87.50 | 98.96 | A |
| Average diastolic | 41.67 | 78.13 | 90.63 | C |

### Comparison of BP classification is important for validation

It is inherently challenging for new blood-pressure measurement devices to meet AAMI and BHS validation criteria when comparisons are made directly with sphygmomanometric absolute values of systolic and diastolic pressures. Device-reference disagreement may reflect error and biological variability in either method; therefore, disagreement should not automatically be attributed entirely to the test device.

If, however, one assumes that ‘classification’ of blood pressure based on repeated sphygmomanometric measurements within an individual is reasonably accurate—since diagnostic cut-offs for hypertension already incorporate the systematic errors inherent in Sphygmomanometry—then comparing BP classification would complement the numerical agreement analysis for validation of a new device.

The 96 participants were classified into 83 Normotensives and 13 hypertensives based on their sphygmomanometric systolic and diastolic pressures (average of 3 recordings) using thresholds of ≥ 140 mmHg systolic or ≥ 90 mmHg diastolic pressures.

Statistics for sphygmomanometric data (average of 3 recordings, M1-M3, all units except N is in mmHg, N is the sample size) are given in Table 4.

**Table 4:** Descriptive statistics of sphygmomanometric pressures.

| **Sphygmomanometry** | **N** | **Mean** | **SD** | **median** | **IQR** | **SEM** | **CV percent** |
| --- | --- | --- | --- | --- | --- | --- | --- |
| **Whole sample** |  |  |  |  |  |  |  |
| Average systolic | 96 | 114 | 14.71 | 111 | 18.00 | 1.50 | 12.89 |
| Average diastolic | 96 | 76 | 11.55 | 75 | 13.50 | 1.18 | 15.21 |
| Avg Pulse Pressure | 96 | 38 | 10.83 | 37 | 14.00 | 1.11 | 28.37 |
| **Normotensives by Reference sphygmomanometry** |  |  |  |  |  |  |  |
| Average systolic | 83 | 110 | 10.66 | 109 | 15.00 | 1.17 | 9.68 |
| Average diastolic | 83 | 73 | 8.15 | 73 | 11.75 | 0.89 | 11.21 |
| Avg Pulse Pressure | 83 | 38 | 10.40 | 36 | 14.50 | 1.14 | 27.72 |
| **Hypertensives by Reference sphygmomanometry** |  |  |  |  |  |  |  |
| Average systolic | 13 | 139 | 12.50 | 140 | 14.25 | 3.47 | 9.00 |
| Average diastolic | 13 | 97 | 8.00 | 98 | 7.50 | 2.22 | 8.28 |
| Avg Pulse Pressure | 13 | 42 | 12.93 | 40 | 13.00 | 3.59 | 30.50 |

Statistics for Plethysmometry data for the same groups shown above for average systolic, diastolic, mean arterial and pulse pressures are given in Table 5:

**Table 5:**
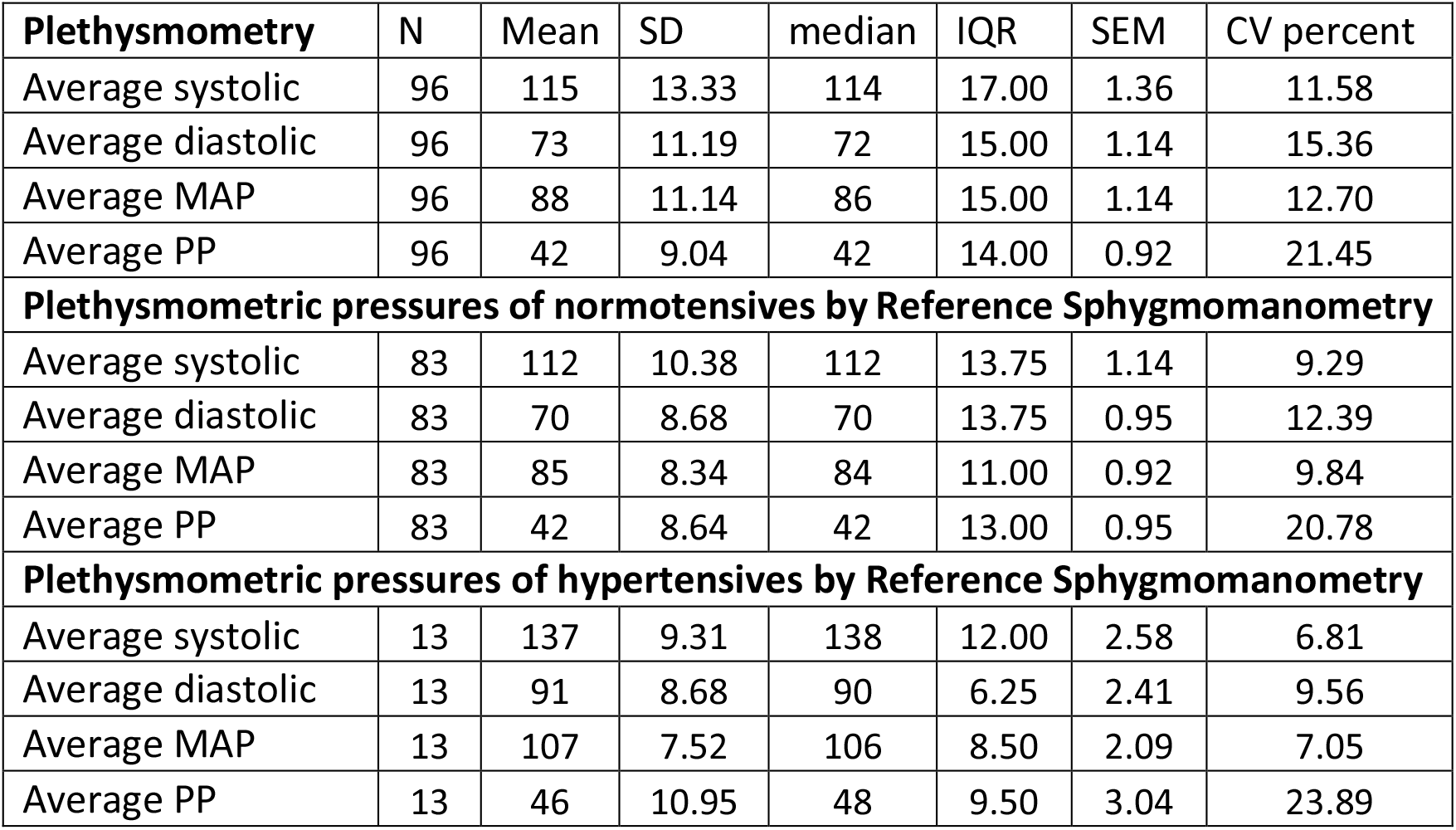
Descriptive statistics of plethysmometric pressures for the same groups as in Table 4.

| **Plethysmometry** | **N** | **Mean** | **SD** | **median** | **IQR** | **SEM** | **CV percent** |
| --- | --- | --- | --- | --- | --- | --- | --- |
| Average systolic | 96 | 115 | 13.33 | 114 | 17.00 | 1.36 | 11.58 |
| Average diastolic | 96 | 73 | 11.19 | 72 | 15.00 | 1.14 | 15.36 |
| Average MAP | 96 | 88 | 11.14 | 86 | 15.00 | 1.14 | 12.70 |
| Average PP | 96 | 42 | 9.04 | 42 | 14.00 | 0.92 | 21.45 |
| **Plethysmometric pressures of normotensives by Reference Sphygmomanometry** |  |  |  |  |  |  |  |
| Average systolic | 83 | 112 | 10.38 | 112 | 13.75 | 1.14 | 9.29 |
| Average diastolic | 83 | 70 | 8.68 | 70 | 13.75 | 0.95 | 12.39 |
| Average MAP | 83 | 85 | 8.34 | 84 | 11.00 | 0.92 | 9.84 |
| Average PP | 83 | 42 | 8.64 | 42 | 13.00 | 0.95 | 20.78 |
| **Plethysmometric pressures of hypertensives by Reference Sphygmomanometry** |  |  |  |  |  |  |  |
| Average systolic | 13 | 137 | 9.31 | 138 | 12.00 | 2.58 | 6.81 |
| Average diastolic | 13 | 91 | 8.68 | 90 | 6.25 | 2.41 | 9.56 |
| Average MAP | 13 | 107 | 7.52 | 106 | 8.50 | 2.09 | 7.05 |
| Average PP | 13 | 46 | 10.95 | 48 | 9.50 | 3.04 | 23.89 |

Receiver operating characteristic (ROC) analysis was performed to assess the ability of plethysmometric systolic pressure, diastolic pressure, and mean arterial pressure to distinguish reference hypertensive from reference non-hypertensive subjects (Figure 4). The ROC curves demonstrate that systolic, diastolic, as well as mean arterial pressures, when used as diagnostic thresholds individually, have AUC > 0.9.

**Figure 4:**
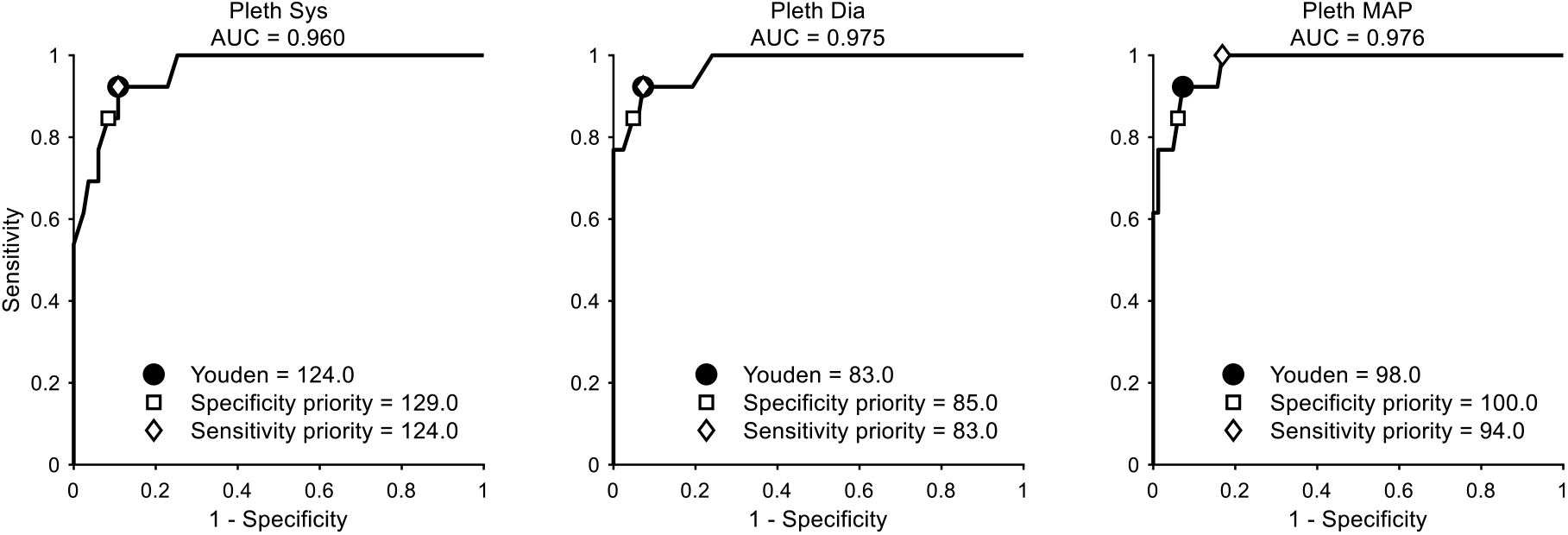
ROC curves assessing the ability of plethysmometric systolic, diastolic and mean arterial pressure thresholds to classify BP equivalent to reference sphygmomanometry.

From the ROC analysis, individual and combined criteria with both sensitivity and specificity exceeding 80% were selected for presentation. Table 6 shows their classification performance relative to reference sphygmomanometry.

**Table 6:** Evaluation of diagnostic thresholds for hypertension diagnosis with plethysmometry, by comparison with reference sphygmomanometry^1^.

| Condition | Concordant positive | Discordant positive | Concordant negative | Discordant negative | Sensitivity | Specificity | Youden index |
| --- | --- | --- | --- | --- | --- | --- | --- |
| Sys ≥ 124 | 12 | 9 | 74 | 1 | 92.31 | 89.16 | 81.46 |
| Dia ≥ 83 | 12 | 6 | 77 | 1 | 92.31 | 92.77 | 85.08 |
| MAP ≥ 98 | 12 | 6 | 77 | 1 | 92.31 | 92.77 | 85.08 |
| Sys ≥ 124 OR Dia ≥ 83 | 13 | 12 | 71 | 0 | 100 | 85.54 | 85.54 |
| Sys ≥ 124 OR MAP ≥ 98 | 12 | 11 | 72 | 1 | 92.31 | 86.75 | 79.05 |
| Dia ≥ 83 OR MAP ≥ 98 | 13 | 8 | 75 | 0 | 100 | 90.36 | 90.36 |
| Sys ≥ 124 OR Dia ≥ 83 OR MAP ≥ 98 | 13 | 13 | 70 | 0 | 100 | 84.34 | 84.34 |
| Sys ≥ 124 AND Dia ≥ 83 | 11 | 3 | 80 | 2 | 84.62 | 96.39 | 81.00 |
| Sys ≥ 124 AND MAP ≥ 98 | 12 | 4 | 79 | 1 | 92.31 | 95.18 | 87.49 |
| Dia ≥ 83 AND MAP ≥ 98 | 11 | 4 | 79 | 2 | 84.62 | 95.18 | 79.80 |
| Sys ≥ 124 AND Dia ≥ 83 AND MAP ≥ 98 | 11 | 3 | 80 | 2 | 84.62 | 96.39 | 81.00 |

Among the evaluated single and combined thresholds, the criterion Dia ≥ 83 mmHg OR MAP≥98 mmHg provided the highest Youden index, with a sensitivity of 100% and specificity of 90.36%. The Youden index was calculated as sensitivity + specificity ™100 because sensitivity and specificity are expressed as percentages.

The terms true positive, false positive, true negative and false negative have been avoided because disagreement with sphygmomanometric classification does not necessarily establish that the plethysmometric classification is erroneous. Instead, concordant positive, discordant positive, concordant negative and discordant negative are used to describe agreement or disagreement with the reference classification.

Although the criterion diastolic pressure ≥83 mmHg OR MAP ≥ 98 mmHg had the highest Youden index, it does not include systolic pressure and may therefore fail to identify isolated systolic hypertension. It should not be interpreted as a universally preferable diagnostic rule.

For screening, where failure to identify possible hypertension is of greater concern, the combined criterion of (systolic pressure ≥ 124 mmHg OR diastolic pressure ≥ 83 mmHg) AND MAP ≥ 98 mmHg may be preferred. This criterion incorporates all three pressure parameters and reported hypertension in 12 of the 13 participants classified as hypertensive by reference sphygmomanometry. There was one discordant negative classification. The criterion also led to five more discordant positive classifications. The corresponding contingency table is shown in Table 7.

**Table 7:** Contingency table for diagnostic performance of Plethysmometry as compared to reference sphygmomanometry with the diagnostic criterion as (Sys ≥124 mmHg OR Dia ≥ 83 mmHg) AND MAP ≥ 98 mmHg.

|  |  | Reference sphygmomanometry |  |
| --- | --- | --- | --- |
|  |  | HT (13) | NT (83) |
| Plethysmometry | HT (17) | 12 | 5 |
|  | NT (79) | 1 | 78 |

The sensitivity of the criterion is 0.923 and specificity, 0.94. The positive and negative predictive values are respectively 0.706 and 0.987. The six participants with discordant classifications require carefully controlled repeat measurements using both methods before their pressure status can be determined. The five discordantly positive participants may represent masked hypertension, whereas the lone discordantly negative participant may represent hypertension that was not identified by the selected plethysmometric thresholds, or it may represent a case of white coat hypertension.

When systolic pressure alone exceeds its threshold, while MAP remains below its threshold, a wide pulse pressure may also be included in the criterion for identifying such cases of isolated systolic hypertension.

The plethysmometric and sphygmomanometric measurements of the five discordantly positive participants are presented in Table 8. The corresponding measurements of the lone discordantly negative participant are presented in Table 9.

**Table 8:** Pressure values of the five discordant positive cases shown in Table 7.

| Plethysmometry-positive discordant cases |  |  |  |  |  |  |  |  | Reference<br>Sphygmomanometry |  |
| --- | --- | --- | --- | --- | --- | --- | --- | --- | --- | --- |
| LS | HS | LD | HD | LMAP | HMAP | Avg Sys | Avg Dia | Avg MAP | Avg Sys<br>(M1 to M3) | Avg Dia<br>(M1 to M3) |
| 120 | 125 | 86 | 88 | 100 | 102 | 123 | 87 | 101 | 109 | 77 |
| 129 | 138 | 79 | 83 | 99 | 101 | 134 | 81 | 100 | 139 | 88 |
| 130 | 140 | 82 | 87 | 104 | 106 | 135 | 85 | 105 | 119 | 80 |
| 133 | 138 | 82 | 87 | 97 | 104 | 136 | 85 | 101 | 126 | 80 |
| 128 | 137 | 81 | 84 | 100 | 101 | 133 | 83 | 101 | 130 | 86 |

**Table 9:** Pressure values for the lone discordantly negative subject in Table 7.

| Plethysmometry |  |  |  |  |  |  |  | Reference<br>sphygmomanometry |  |
| --- | --- | --- | --- | --- | --- | --- | --- | --- | --- |
| LS | HS | LD | HD | LMAP | HMAP | Avg Sys | Avg Dia | Avg Sys | Avg Dia |
| 116 | 119 | 81 | 84 | 91 | 96 | 118 | 83 | 115 | 94 |

Individual sphygmomanometric readings for the five discordantly positive participants and the lone discordantly negative participant are shown in Tables 10 and 11 respectively.

**Table 10:** Reference sphygmomanometric pressures for the four repeats (M0 to M3) for the discordantly classified cases shown in Table 8.

| Average of Observer 1 and 2 |  |  |  |  |  |  |  |  |  |  |  |
| --- | --- | --- | --- | --- | --- | --- | --- | --- | --- | --- | --- |
| M0 |  | M1 |  | M2 |  | M3 |  | Average of M0 to M3 |  | Average of M1 to M3 |  |
| 106 | 71 | 108 | 80 | 109 | 74 | 109 | 77 | 108 | 76 | 109 | 77 |
| 148 | 88 | 137 | 86 | 140 | 93 | 140 | 85 | 141 | 88 | 139 | 88 |
| 140 | 81 | 122 | 81 | 118 | 73 | 118 | 86 | 125 | 80 | 119 | 80 |
| 135 | 91 | 133 | 85 | 122 | 80 | 122 | 76 | 128 | 83 | 126 | 80 |
| 128 | 65 | 134 | 84 | 124 | 88 | 132 | 86 | 130 | 81 | 130 | 86 |

**Table 11:** Reference sphygmomanometric pressures for the four repeats (M0 to M3) for the discordantly classified case shown in table 9.

| Average of Observer 1 and 2 |  |  |  |  |  |  |  |  |  |  |  |
| --- | --- | --- | --- | --- | --- | --- | --- | --- | --- | --- | --- |
| M0 |  | M1 |  | M2 |  | M3 |  | Average of M0 to M3 |  | Average of M1 to M3 |  |
| 122 | 96 | 116 | 97 | 118 | 96 | 112 | 89 | 117 | 95 | 115 | 94 |

The average of M1 to M3 has been used for diagnostic classification in this study.

### Relationship between the ROC-derived thresholds and the observed pressure distributions

Table 12 presents selected descriptive quantities derived from the distributions of plethysmometric pressures in the whole sample and in the groups classified as hypertensive or not hypertensive by reference sphygmomanometry. These values are provided only to describe the position of the ROC-derived thresholds relative to the observed pressure distributions; they were not used to define the diagnostic thresholds.

**Table 12:** Relationship of the ROC-derived thresholds to selected descriptive values from the pressure distributions.

| Pressure parameter (mmHg) | Plethysmometry |  |  |  |  | Sphygmomanometry |  |  |
| --- | --- | --- | --- | --- | --- | --- | --- | --- |
|  | ROC-derived threshold (for sensitivity) | ROC-derived threshold (for specificity) | Whole Sample Mean + SD | NT Mean + 1.5* SD | HT Mean - SD | Whole Sample Mean + SD | NT Mean + 1.5*SD | HT Mean - SD |
| Avg Sys | 124 | 128 | 128 | 127 | 127 | 129 | 126 | 126 |
| Avg Dia | 83 | 85 | 84 | 83 | 82 | 87 | 85 | 89 |
| Avg MAP | 98 | 100 | 99 | 97 | 99 | - | - | - |

It is seen from Table 12 that the ROC-derived systolic threshold for specificity is closer to whole sample mean + SD, Normotensive mean + 1.5*SD and Hypertensive mean minus SD. If the plethysmometric diagnostic criterion used values from the third column above, ie, the specificity thresholds, the discordant positives remained the same, but there was one other discordant negative as compared to Table 7.

Raw traces corresponding to the participant in the third row of Table 8 are shown in Figure 5.

**Figure 5:**
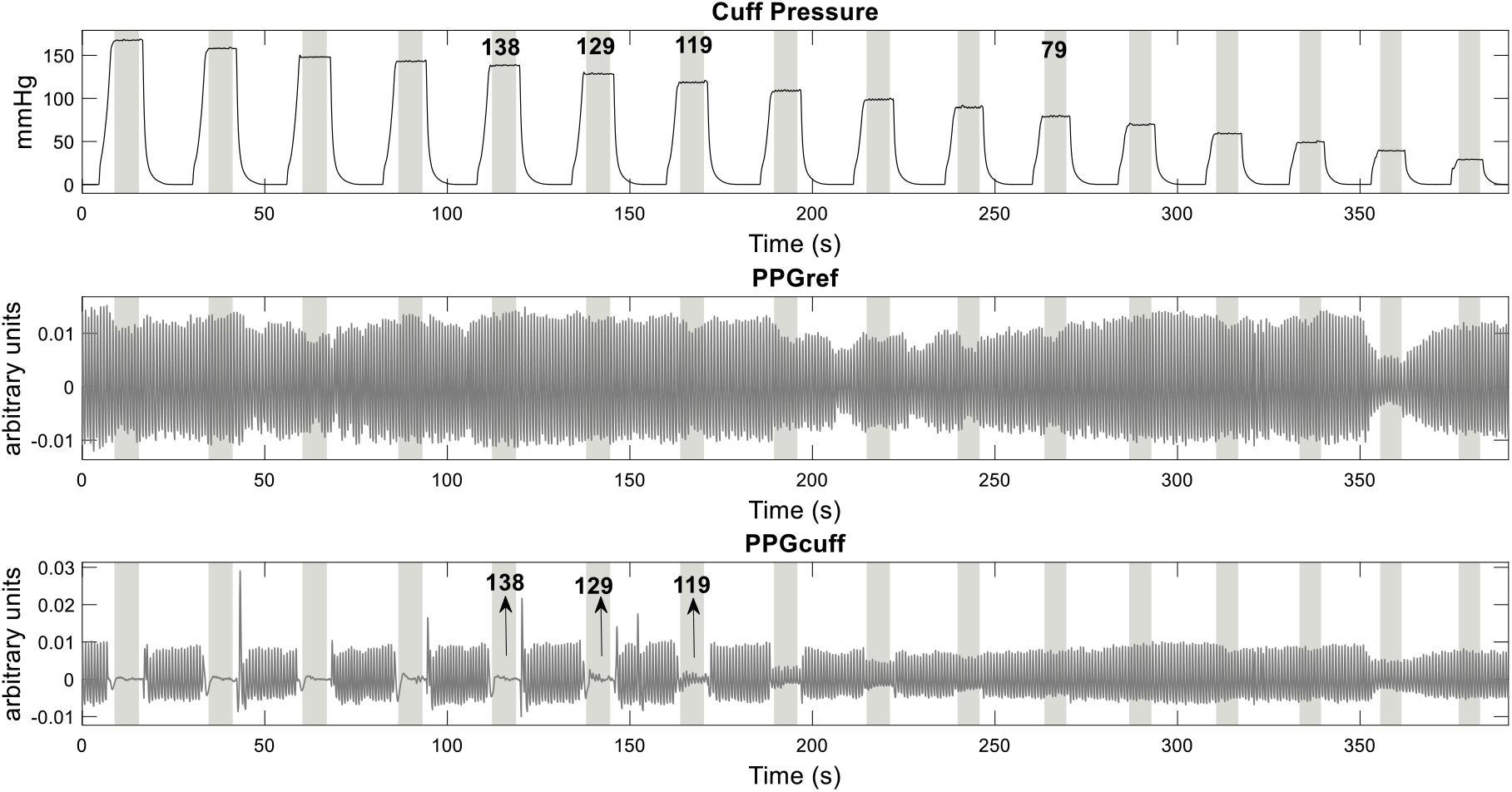
Raw plethysmometric tracings from a participant with discordant positive classification.

Sphygmomanometric systolic estimation for the subject whose data is found in Figure 5 is 119 mmHg. Note that all pulses are present at cuff pressures of 119 and some at 129. Only at 138 mmHg, all pulses are absent. Our report for systolic pressure in this subject is 130-140 mmHg.

## Discussion

In the present study, Sphygmomanometry was performed under highly controlled conditions—in a sound-proof room with simultaneous video recording of the mercury column and audio recording of Korotkoff sounds. Each subject underwent four measurements, two before and two after Plethysmometry. Two observers independently reported systolic and diastolic pressures; if their readings differed by more than 2 mmHg, a third observer acted as arbiter. Despite these precautions, Korotkoff sounds did not always appear and disappear precisely at systolic and diastolic pressures, suggesting that additional factors such as arterial wall vibration influence sound generation.

Consequently, when a new device is compared directly with Sphygmomanometry in terms of absolute pressures, device-reference disagreement may reflect error and biological variability in either method; therefore, disagreement should not automatically be attributed entirely to error in the test device.

Sphygmomanometry is traditionally considered the reference standard for validating new blood pressure (BP) measuring devices. Because sphygmomanometry relies on auscultation, which is subject to physiological and observer variability, validating a device which reports a pressure range rather than a single value, by comparison based on absolute single-value differences may not capture all aspects of its performance.

In this study, systolic pressure met the numerical requirements of AAMI/ESH/ISO Criterion 1 for mean difference and standard deviation, but the Bland–Altman limits of agreement were relatively wide, extending from ™11.94 to 13.87 mmHg. Thus, small mean differences and fulfilment of the Criterion 1 numerical requirements do not imply close agreement with the reference method for every individual measurement. Differences of this magnitude could alter hypertension classification in participants whose pressures lie near a diagnostic threshold. These findings support evaluating diagnostic sensitivity and specificity when BP devices are used to classify hypertension, in addition to assessing population-level numerical agreement with the reference method.

Given that sphygmomanometry has been in clinical use for more than a century and is employed by millions of healthcare professionals worldwide, the diagnostic thresholds for hypertension are necessarily comprehensive, having implicitly accommodated the probable limitations of this method.

Numerical agreement and diagnostic classification address related but distinct aspects of performance. Agreement analysis assesses the magnitude of device-reference differences, whereas classification analysis assesses whether those differences change the clinical category of the estimated pressure. We therefore evaluated both outcomes. The underlying assumption is that averaged pressures from multiple sphygmomanometric measurements in the same individual provide a reasonably accurate classification of hypertensive versus normotensive status.

Using the selected exploratory criterion, plethysmometry achieved sensitivity and specificity above 0.9 relative to the reference sphygmomanometric classification in this cohort. A single automated plethysmometric recording showed promising classification agreement with the reference category derived from multiple sphygmomanometric measurements that require trained observers and ideal acoustic conditions. However, the thresholds identified in this study should be considered exploratory.

As stated in the accompanying paper, there are three broad approaches to deriving classification thresholds: distribution-based, reference-comparison-based and outcome-based. Although outcome-based thresholds are ultimately preferable, reference-comparison and ROC-derived thresholds may be used provisionally until longitudinal outcome data become available.

Masked HT can only be detected through out-of-office monitoring, such as home BP recordings or 24-hour ambulatory BP monitoring (ABPM). Currently, subjects with normal office BP are rarely evaluated further; hence, masked HT remains undiagnosed. Whether plethysmometry can identify individuals with masked hypertension should be evaluated prospectively against ABPM or structured HBPM.

Interestingly, the mean systolic and diastolic pressures for the whole sample and for the normotensives for both methods are close.

## Future Directions

Further studies should compare plethysmometric pressures with intra-aortic and intra-arterial measurements to confirm that plethysmometric values correspond to aortic/arterial pressures. The plethysmometric diagnostic thresholds and combined classification rules must be confirmed or refined in larger, independent datasets. The systolic and diastolic variability parameters (difference between upper and lower estimates) provided by the method must be evaluated for functional significance.

## Conclusion

In exploratory analysis, plethysmometric systolic, diastolic and mean arterial pressure thresholds, used singly and in combination, showed promising discrimination of sphygmomanometrically defined hypertension. The criterion diastolic pressure ≥ 83 mmHg OR MAP ≥98 mmHg provided the highest Youden index. However, the criterion used in this study includes systolic pressure parameter as well to reduce the risk of missing isolated systolic hypertension. Because these thresholds were derived and evaluated in the same small cohort, they require independent validation using a locked algorithm and prespecified criteria.

## ACKNOWLEDGEMENTS

The authors thank the Dept. of Physiology, Christian Medical College, Vellore for enabling the study and data collection.

## ETHICS

This study was approved by the Institutional Review Board of Christian Medical College, Vellore, IRB no: 14388, 22/12/2021

## DATA AVAILABILITY STATEMENT

The datasets generated during and/or analysed during the current study are available from the corresponding author on reasonable request.

## Footnotes

1 Sys, Dia and MAP denote plethysmometric midpoint systolic, diastolic and mean arterial pressures, respectively. Candidate thresholds were identified from ROC analysis and evaluated in the same cohort; the findings are therefore exploratory.

